# Smart AI-Powered Machine Learning Risk Assessment for Early Osteoporosis Detection for Women’s Bone Health

**DOI:** 10.64898/2026.05.31.26354550

**Authors:** Vahid Monfared

## Abstract

Osteoporosis is often called a “silent disease” because it progresses without symptoms until a fracture occurs, posing a serious, yet frequently overlooked, threat to women’s health. In response to the pressing need for early detection, we introduce OsteoInsight, an intelligent, AI-powered web application designed to assess osteoporosis risk with both clinical accuracy and interpretability. Built on a Random Forest classifier trained on over 2000 women’s health records, our model incorporates a wide range of domain-informed features, including hormonal history, lifestyle factors, reproductive health, and conditions affecting bone health. Despite an imbalanced dataset, with around 75% of cases being osteoporosis-positive, the model achieved encouraging results: 71.81% accuracy, an F1-score of 0.79, and an AUC-ROC of 0.78. SHAP analysis highlighted age, BMI, and menstrual history as key predictors, offering transparent insights into the model’s reasoning. Additional contributors like fracture history, signs of low estrogen, and lactation duration were also found to be significant, enriching the interpretability of predictions.

These insights are seamlessly integrated into OsteoInsight’s user interface, making risk assessments not only accessible but also understandable for both clinicians and users. Our findings underscore the potential of AI-driven tools to enhance early screening and enable personalized risk profiling, empowering women and healthcare providers to take proactive steps in osteoporosis prevention.

## 1. Introduction

Osteoporosis, often referred to as the “silent thief,” gradually weakens bones without warning signs until a fracture occurs. It disproportionately affects women, especially during and after menopause, due to hormonal shifts that accelerate bone loss. Despite the known consequences, pain, loss of mobility, and increased mortality after hip fractures, screening remains underutilized in many regions. Dual-energy X-ray absorptiometry (DXA) scans, the gold standard for diagnosis, are expensive and limited in accessibility. Consequently, there’s a pressing need for alternative, scalable, and early detection methods that can identify at-risk women before fractures occur.

Early efforts in osteoporosis screening produced relatively simple clinical tools. For example, the Osteoporosis Risk Assessment Instrument (ORAI) was developed by Cadarette et al. in a Canadian cohort using age, weight and current estrogen use as predictors; it achieved very high sensitivity but modest specificity [1]. Subsequent work by Gourlay et al. evaluated ORAI and similar tools in older U.S. women and found only fair discrimination (AUC ≈ 0.7) [2]. These tools were useful but lacked deeper personalization and generalizability. With the advent of machine learning (ML), researchers moved to more data-driven models. For instance, Suh et al. developed an interpretable deep-learning model for osteoporosis screening with clinical features [3]. Cha et al. used ML feature-selection to build risk-prediction models and showed that ML can select non-traditional predictors effectively [4]. A systematic review by Wu et al. found that ML has favorable predictive performance for fracture risk in osteoporosis but noted poor external validation across settings [5]. The national-scale study by Tu et al. employed chronic disease data from Germany (n ≈ 10 000) and built a stacker ML model with age, gender, lipid disorders, cancer, COPD and other features, achieving strong results and showing promise for broader screening [6]. More recently, Si et al. used NHANES data and found LightGBM achieved an AUC ≈ 0.97 for osteoporosis identification in a large U.S. sample [7]. Another effort by Carvalho et al. specifically built a ML model excluding DXA inputs, aiming at more accessible tools in limited-resource settings [8]. The 2025 work by Je et al. focused on Korean women (n ≈ 4 199) using ML versus traditional tools and confirmed that ML can outperform conventional instruments [9]. Yet, the latest study by Elias et al. emphasized explainability (XAI) in osteoporosis risk models, integrating SHAP/LIME so clinicians can understand model reasoning rather than operate a black box [10]. From this narrative we see a clear progression, from simple scoring tools through ML models to interpretable AI, but also considerable gaps: many models rely on specific populations (ethnicities, geographies), many embed DXA or imaging inputs (limiting scalability), and few integrate interpretability with clinically meaningful outputs in Web-based tools usable in real-world settings. This gap underscores the need for a women-centric, geographically diverse, interpretable and deployable risk-assessment tool built on real-world hospital data. To fill this void, our study introduces OsteoInsight, an AI-powered web application built on hospital data. Another main point is in obtaining important features and orders of those, and impact of features levels on target which is very highlighted and presented very useful information for women to early detection and prevention of osteoporosis risks (and a great web application). Our model is trained on over 2000 women’s health records incorporating features often excluded from conventional risk models: hormonal history, menstrual and reproductive data, fracture history, and lifestyle factors. With a Random Forest backbone and explainability powered by SHAP values, our system not only predicts risk with solid accuracy (71.81%, precision: 0.88, F1-score of around 0.8, AUC-ROC of 0.78) but also provides transparency into *why* a prediction was made, empowering both clinicians and users. Unlike black-box systems, OsteoInsight visualizes key drivers of risk such as BMI, estrogen-related factors, and lactation history, offering a truly personalized snapshot.

Its web interface makes it accessible for clinical use and public health screening, especially in underrepresented settings. In summary, our tool bridges several known gaps in osteoporosis risk prediction and demonstrates how region-specific, explainable AI can have real-world healthcare impact, paving the way for more proactive and equitable bone health management.

The web application and GitHub resource are available here https://osteo-insight-hub.lovable.app/ and GitHub: https://github.com/VahidMonfared/AI-Powered-Osteoporosis-Detection respectively.

## 2. Methodology

### 2.1 Dataset Overview and Feature Composition

Our study utilized a real-world clinical dataset comprising over 2000 records of women, designed specifically for osteoporosis risk prediction. Each record consisted of 22 attributes; 21 input features and one target variable, carefully selected to reflect medical, reproductive, and lifestyle dimensions relevant to women’s bone health. These features captured data on age, BMI, menstrual and reproductive history, hormone-related indicators, fracture history, physical activity, nutritional intake (e.g., calcium, vitamin D, dairy), and past medical conditions. This diverse and clinically grounded dataset allowed us to build a multidimensional view of osteoporosis risk, with particular relevance to underrepresented healthcare populations. Among the features, four were continuous variables: *Age, BMI, Gravidity* (number of pregnancies), and *Parity* (number of births). The remaining 17 were categorical, spanning clinical signs, menstrual patterns, estrogen-related symptoms, surgical history, lifestyle factors, and chronic health conditions. Each categorical feature was transformed using One-Hot Encoding (OHE) to retain interpretability and prepare for SHAP-based analysis later. The target variable, Osteoporosis_Total, was a binary label: ‘1’ indicated a confirmed osteoporosis diagnosis, while ‘2’ represented absence of osteoporosis. For classification purposes, we internally recoded this as a binary outcome: 1 = At Risk, 0 = Not at Risk (see Fig.1).

**Figure 1.**
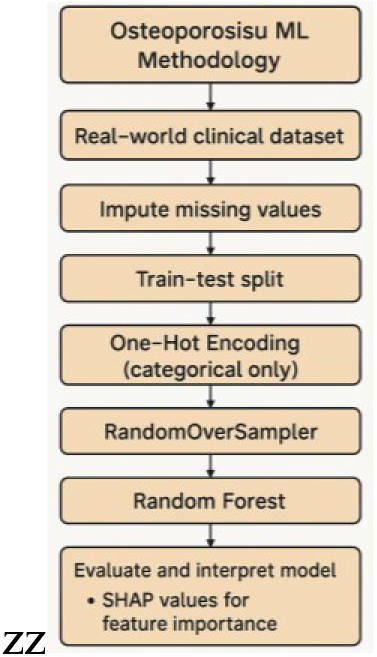
Flowchart and problem solution steps

### 2.2 Preprocessing and Data Imputation

Given the mixed nature of data (categorical and numerical), we employed a tailored imputation strategy (statistical and AI imputations). For continuous features like *Age* and *BMI*, we used iterative imputation to preserve inter-variable relationships. For discrete numerical features such as *Gravidity* and *Parity*, median imputation followed by rounding ensured clinically meaningful values. Categorical features were imputed using the most frequent value method, maintaining data integrity without introducing artificial distributions. Following imputation, no samples were excluded; the full over 2000 records were retained for training and evaluation, ensuring model robustness and generalizability.

### 2.3 Modeling Pipeline and Training

To address the significant class imbalance, with ∼75% of cases being osteoporosis-positive, we integrated RandomOverSampler within our modeling pipeline. This technique synthetically balanced the training set, preventing bias toward the dominant class. We selected the Random Forest (RF) classifier for its performance, interpretability, and adaptability to tabular medical data. The RF model was fine-tuned through an extensive grid search, testing over 400 hyperparameter configurations using 10-fold cross-validation. The best-performing model including, 400 decision trees, maximum depth of 16, “sqrt” strategy for feature selection, optimized thresholds for node splits and leaf sizes.

### 2.4 Evaluation, Explainability, and Deployment

Our final model achieved acceptable performance,

- Training set: Accuracy = 78.1%, F1-score = 0.83, AUC-ROC = 0.90
- Test set: Accuracy = 71.8%, F1-score = 0.80, AUC-ROC = 0.77

These results show the model’s strong predictive capability, even when faced with real-world class imbalance. To ensure transparency, we applied SHAP (SHapley Additive exPlanations) for both global feature importance and per-feature contribution analysis. Key influencers identified included *Age, BMI, Menstrual History, Fracture History*, and *Low Estrogen Symptoms*. SHAP not only supported model interpretability but also enabled clinicians to understand why certain individuals were classified as high-risk. All artifacts preprocessed data, trained model, performance metrics, ROC curves, confusion matrices, and SHAP visualizations, were exported and integrated into our web-based tool, OsteoInsight. The tool is user-friendly and designed for real-time risk prediction. It bridges the gap between AI and clinical practice, helping physicians and patients engage in informed, early-stage osteoporosis screening.

### 2.5 Full List of Features (Inputs)

Here, there are all features such as Age_Year – Age of the individual (in years), BMI_kg_per_m2 –Body Mass Index, Gravidity – Number of pregnancies, Parity – Number of births, Occupation – Employment status, Clinical_Signs – Presence of musculoskeletal symptoms, Fracture_History – History of fractures, Age_Last_Delivery_Year – Time since last delivery, Age_of_Menark_Year – Age at menarche, Duration_Lactation_Year – Total lactation duration, Mestural_History – Type of menstrual history, Hysterectomy – History of hysterectomy, Ovarian_Cystectomy_Oophorectomy –History of ovarian surgeries, Immobilization_History – History of prolonged immobilization, Low_Estrogen_Signs – Symptoms of estrogen deficiency, Infertility_History – History of infertility, Calsium_VitamineD_Supplement – Use of supplements, Dairy_Consumption – Dairy intake level, Physical_Activity – Frequency of physical activity, Medication – Use of risk-related medications, Past_Medical_History – Existing comorbidities (e.g., diabetes, thyroid disorders) Note that, as an example, target output is “Osteoporosis_Total” as a Binary outcome, like 1 = Presence of osteoporosis (At Risk), 2 = No osteoporosis (Not at Risk); Internally encoded as: 1 = At Risk, 0 = Not at Risk.

### 2.6 Web Application Questions (User Interface Mapping)

Each input feature was turned into a clear, intuitive question in the OsteoInsight web application are, What is your age (in years)? → *Age_Year;* What is your Body Mass Index (BMI in kg/m^2^)? → *BMI_kg_per_m2;* How many times have you been pregnant? → *Gravidity;* How many births have you had? → *Parity;* What is your occupation? → *Occupation;* Do you have any clinical signs? → *Clinical_Signs;* Have you ever had a fracture? → *Fracture_History;* How many years ago was your last delivery? → *Age_Last_Delivery_Year;* At what age did you have your first period (menarche)? → *Age_of_Menark_Year;* How long have you breastfed in total? → *Duration_Lactation_Year;* What is your menstrual history? → *Mestural_History;* Have you had a hysterectomy? → *Hysterectomy;* Have you had ovarian surgeries? → *Ovarian_Cystectomy_Oophorectomy;* Do you have a history of immobilization? → *Immobilization_History;* Do you have low estrogen symptoms? → *Low_Estrogen_Signs;* Do you have a history of infertility? → *Infertility_History;* Do you take calcium or vitamin D supplements? → *Calsium_VitamineD_Supplement;* How much dairy do you consume? → *Dairy_Consumption;* How often do you exercise? → *Physical_Activity;* Are you taking any medications? → *Medication;* Do you have any past medical conditions? → *Past_Medical_History*.

### 2.7 Deployment and Web Application

A research demonstration web application was developed to provide an interactive interface for the proposed osteoporosis detection system.

he application is available at: https://osteo-insight-hub.lovable.app/. This tool is intended for research and educational demonstration only and is not approved for clinical diagnosis or treatment decisions.

## 3. Results and Discussions

The model demonstrated robust performance, achieving the following metrics on the test set. Following comprehensive preprocessing and imputation of our dataset comprising over 2000 records (originally with 23 features, reduced to 22 after processing), we observed a significant class imbalance: approximately 75% of the cases were osteoporosis-positive. After handling missing data using median and iterative imputation for numerical features and the most frequent strategy for categorical ones, we trained a Random Forest model with hyperparameters optimized through a 10-fold cross-validated randomized grid search (400 configurations). The final model demonstrated strong performance: on the training set, it achieved 78.1% accuracy, 0.8371 F1-score, and an impressive 0.8988 ROC-AUC. On the test set, despite the imbalance, it maintained robust generalizability with 71.8% accuracy, 0.7927 F1-score, and 0.7760 ROC-AUC. These results confirm the model’s capacity to identify at-risk individuals with clinically acceptable precision and recall. All artifacts including the balanced dataset, trained model, and evaluation metrics were saved for deployment in the OsteoInsight application. The confusion matrices for both training and testing indicate a high true positive rate. On the training set, the model correctly classified 1,182 positive cases and 457 negatives. The ROC curve for the test set also shows an AUC of 0.78, indicating good discriminatory power (Figures 2-9).

**Figure 2.**
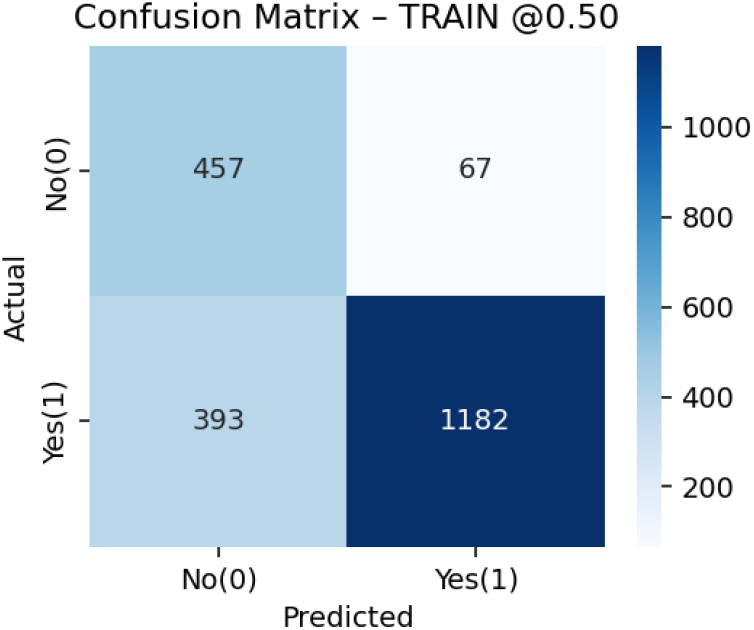
Confusion Matrix – Training Set

Figures 2 and 3 show the confusion matrices for the training and test sets at a 0.50 threshold. The model performed well on the training set, correctly identifying most cases while keeping false positives low. On the test set, it maintained strong recall and precision, confirming its ability to generalize. Despite a few missed or wrongly flagged cases, the results demonstrate the model’s practical value as a reliable screening tool for osteoporosis risk.

**Figure 3.**
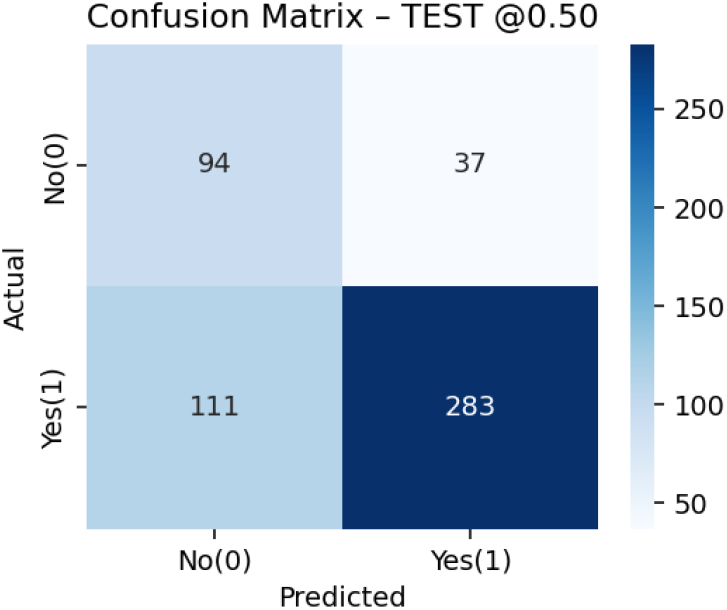
Confusion Matrix – Test Set

Figures 4 and 5 illustrate the ROC curves for the training and test sets, respectively. The model achieved an impressive AUC of 0.90 on the training set, indicating high discriminatory power in distinguishing between individuals at risk and not at risk of osteoporosis. On the unseen test set, the AUC remained strong at 0.78, reflecting good generalization and predictive ability. These curves confirm that the classifier performs well across various thresholds, reinforcing its reliability as a supportive tool for early osteoporosis risk screening.

**Figure 4.**
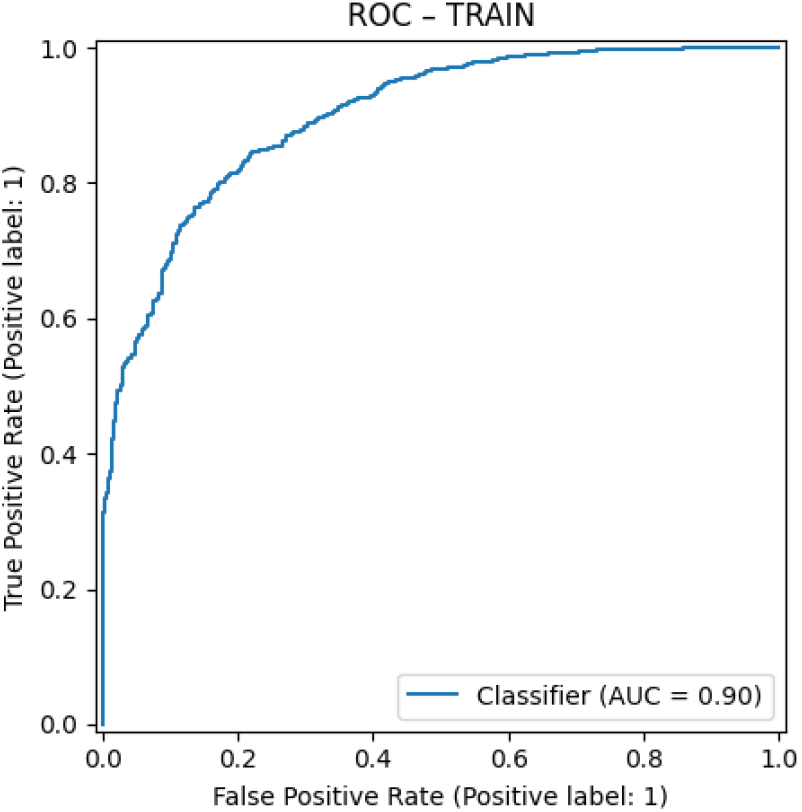
ROC Curve – Training Set

**Figure 5.**
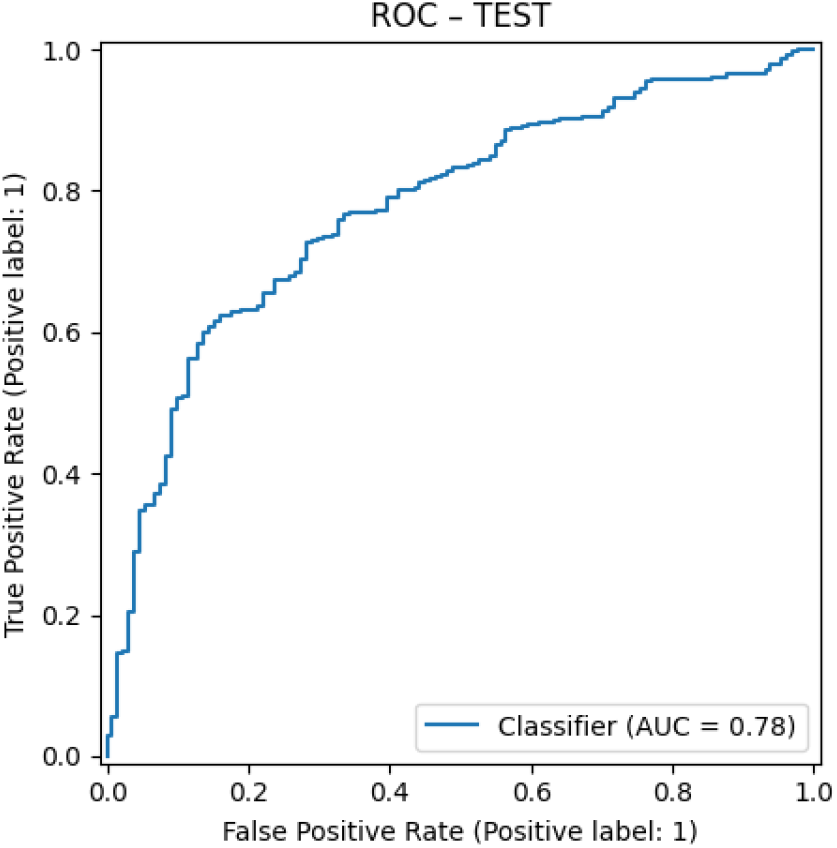
ROC Curve – Test Set

## 4. Feature Importance and Interpretability

Random Forest feature importance revealed that the most significant predictors were Age, BMI, and Menstrual History. SHAP analysis further validated these findings, highlighting the contributions of fracture history, estrogen levels, and reproductive factors. Figures below illustrate global and level-wise SHAP feature importance, showcasing the directionality and magnitude of each factor’s impact on the model’s output.

Figures 6 and 7 highlight the subtle yet important differences in how feature importance is interpreted. In the Random Forest model, Age, BMI, and Menstrual History were the top predictors, reflecting how frequently these features guided the model’s decision paths. However, SHAP analysis reshuffled this order, placing Menstrual History as the most influential, followed by Age and BMI. This suggests that while Age and BMI are structurally important within the model, Menstrual History plays a more decisive role in shifting individual risk predictions. Such divergence emphasizes the value of explainable AI: SHAP uncovers the personalized impact of features that might otherwise be underestimated in traditional importance metrics. Together, these findings underscore the clinical relevance of reproductive history in osteoporosis risk—and reinforce our model’s strength in delivering both accurate and interpretable results.

**Figure 6.**
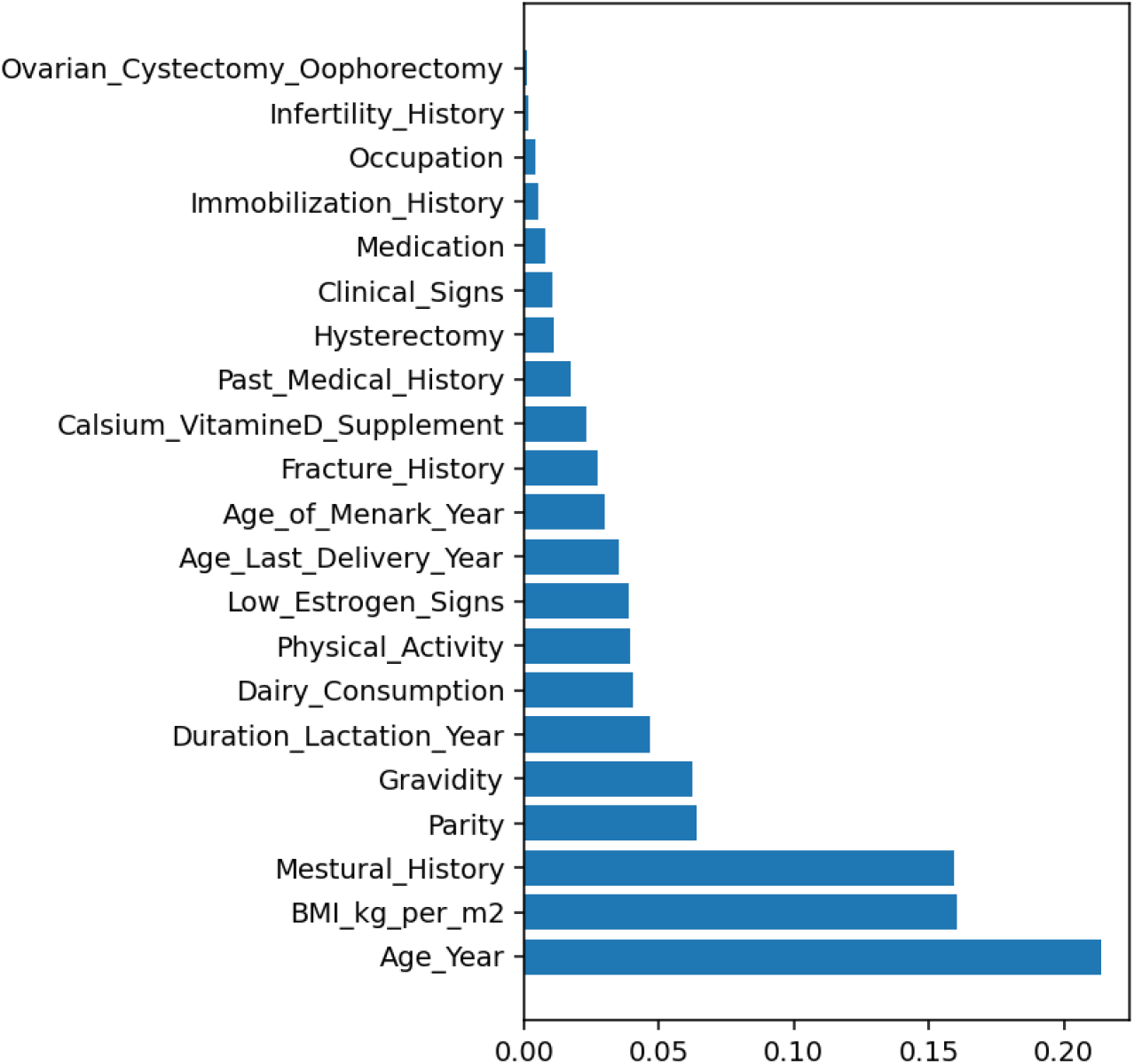
Random Forest Feature Importance

**Figure 7.**
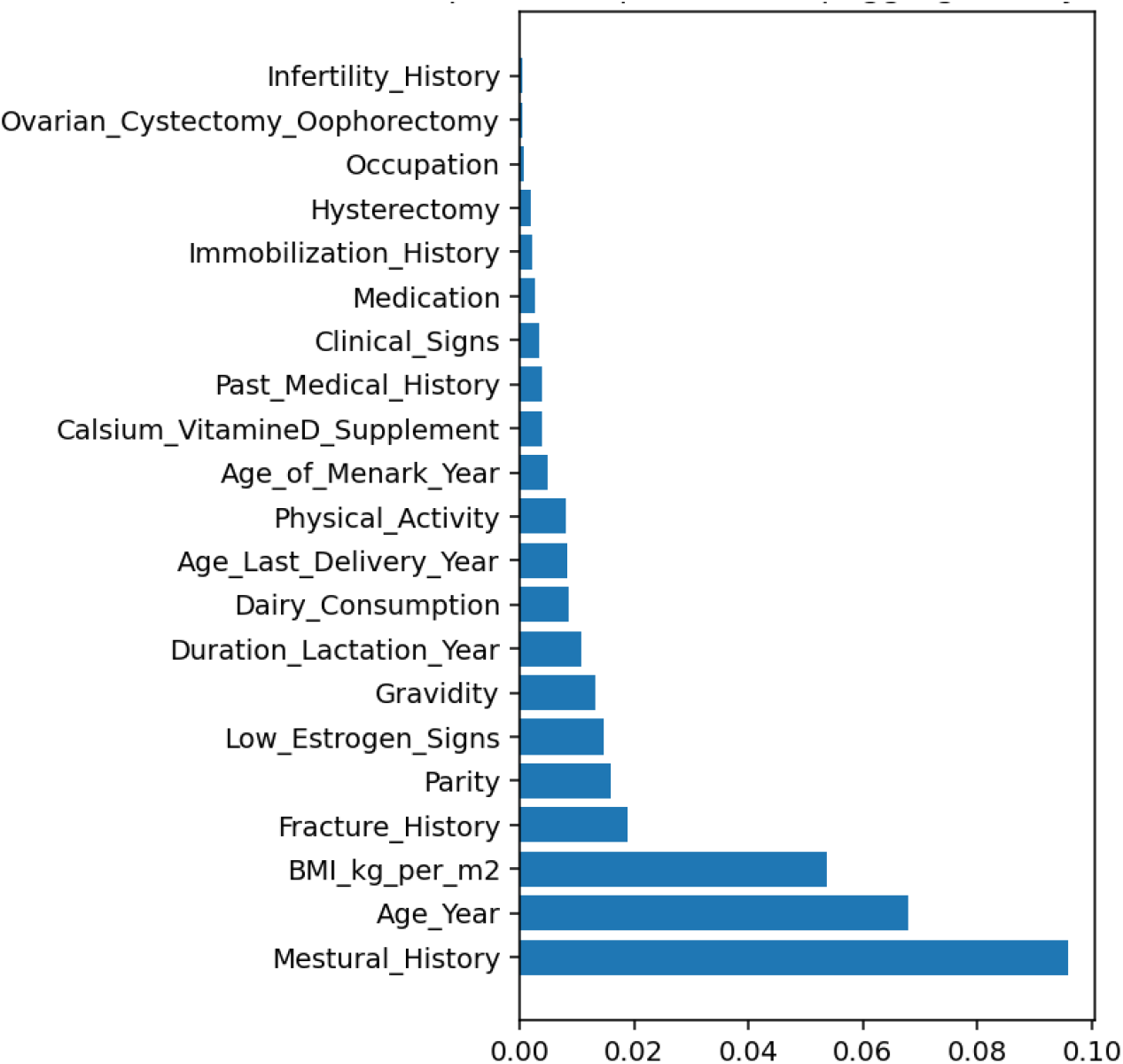
SHAP Global Feature Importance

### Positive Impact on Osteoporosis Risk (↑ risk)

These levels increase the model’s predicted risk of osteoporosis (see Figure 8):

- Menstrual History: Postmenopausal, Strongest positive contributor; postmenopausal women are at significantly higher risk.
- Fracture History: Yes, Prior fractures are a well-known risk factor and are strongly linked to higher osteoporosis likelihood.
- No Previous Pregnancy, Women who have never been pregnant show increased risk, potentially due to hormonal and bone metabolism patterns.
- BMI (Higher), Surprisingly, in this dataset, higher BMI shows a slight risk increase, possibly reflecting comorbidities or fat distribution.
- No Dairy Consumption, Lack of dietary calcium intake is associated with higher bone fragility.
- Menstrual History: Menometrorrhagia / Oligomenorrhea — Irregular or heavy bleeding may indicate hormonal imbalances linked to bone loss.
- Thyroxine Use, Hormone medication, such as thyroxine, is linked to elevated risk, possibly due to accelerated bone turnover.
- No Clinical Signs, Even in the absence of symptoms, some underlying risk factors may be present.
- Hypothyroidism, This condition negatively affects bone density and increases risk.
- Older Age, As expected, age is one of the most significant risk amplifiers.
- Immobilization: Yes, Limited movement contributes to bone demineralization.
- Hot Flashes / Low Estrogen Signs, Reflect estrogen deficiency, directly linked to osteoporosis.
- Premature Menopause, Earlier loss of estrogen protection is a major risk factor.
- Long Lactation Duration, Extended breastfeeding can sometimes deplete maternal calcium stores.
- Very Late Menarche (>16 years), Delayed puberty may be tied to weaker peak bone mass.
- No Estrogen Supplementation, Lack of hormone therapy in postmenopausal women may leave bones unprotected.
- Yes to Infertility / Diabetes / Rheumatoid Arthritis, These medical histories are all linked to lower bone health.
- Hot Flashes + Urinary Symptoms, Combined estrogen deficiency signs are stronger predictors of risk.

**Figure 8.**
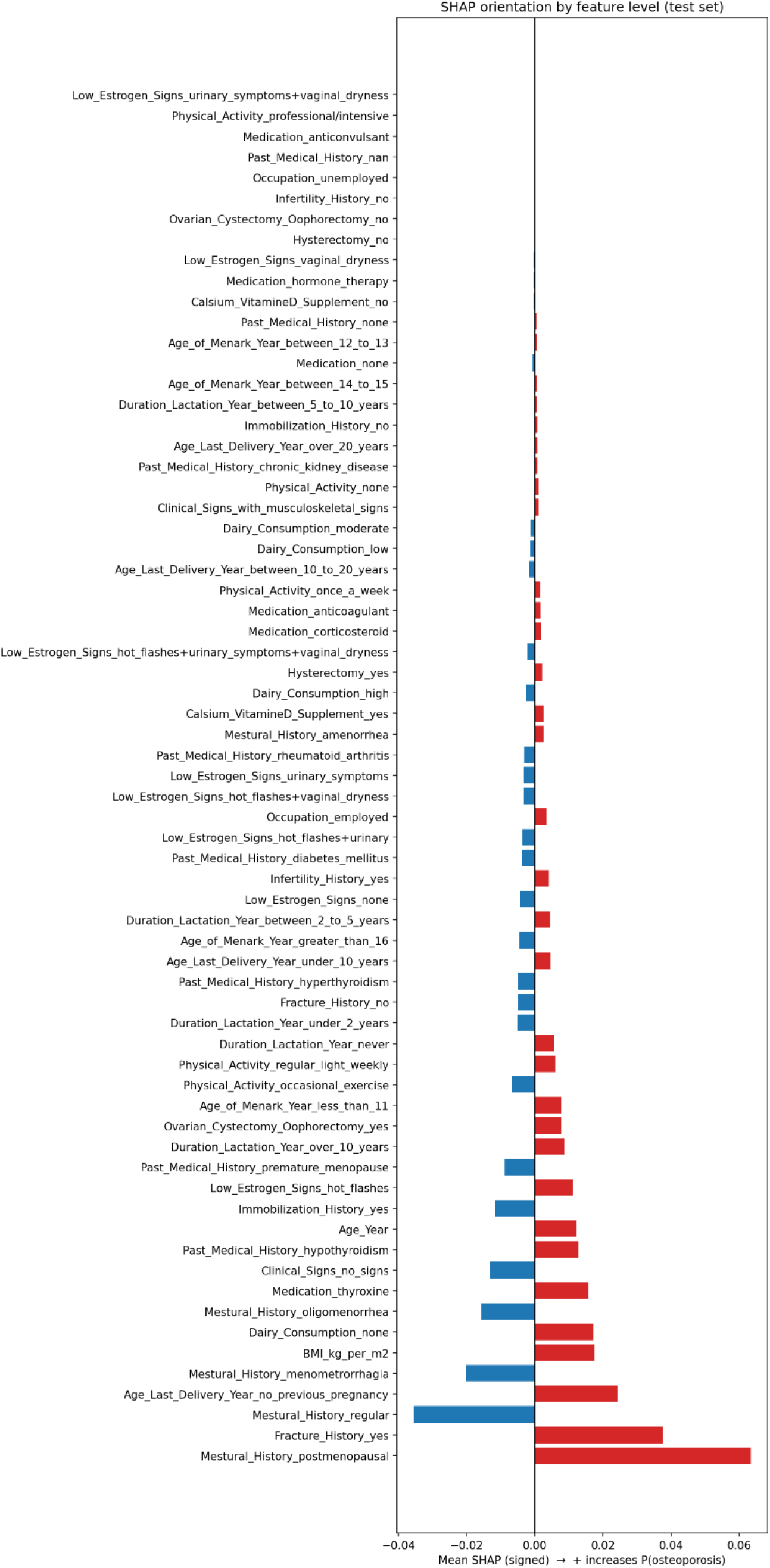
SHAP Orientation by Feature – Test Set

### Negative Impact on Osteoporosis Risk (↓ risk)

These levels decrease the model’s predicted osteoporosis risk:

- Menstrual History: Regular, Indicates hormonal balance, protective for bones.
- No Fracture History, Expectedly, absence of fractures is linked to lower risk.
- No Previous Medical Conditions, Particularly no chronic diseases means fewer indirect risk factors.
- Dairy Consumption: High / Moderate, Regular intake of calcium-rich foods supports bone health.
- Calcium/Vitamin D Supplementation: Yes, Directly supports bone density and reduces risk.
- Physical Activity: Regular or Intensive, Bone-strengthening effects are well-documented.
- Hysterectomy: No, Retaining reproductive organs may preserve hormonal balance.
- Hormone Therapy: Yes, Estrogen replacement helps maintain bone strength in postmenopausal women.
- Early Menarche (12–13 years), Early onset of menstruation may be linked to higher lifetime estrogen exposure, helping bones.

### Neutral or Mixed Effects

Some levels had minimal impact or varied effects:

- Occupation (Unemployed vs. Employed), Socioeconomic and activity level implications, but not strongly directional.
- Medication: Anticoagulants / Corticosteroids / Anticonvulsants, Known risks, but effects in this dataset are relatively mild.
- Mild Physical Activity / Occasional Exercise, Help but less impactful than regular or intensive activity.
- Various Lactation Durations (under 2 to 10 years), Effects vary; mild lactation may be protective, while extended periods may deplete resources.
- Delivery Timing (10–20 years ago vs. recently), Not strongly predictive alone.
- Low Estrogen Symptoms: Mixed combinations, Directionality varies with symptom severity and combinations.

This per-level SHAP analysis gives us a nuanced view of how each health, reproductive, and lifestyle factor influences osteoporosis risk. Some levels clearly signal higher risk, such as postmenopausal state, prior fractures, and lack of calcium intake, while others, like physical activity and regular menstruation, act as protectors. By identifying these level-specific effects, our model helps clinicians and patients make informed, personalized decisions for early intervention and preventive care.

Our model shows strong potential in accurately identifying women at risk for osteoporosis using explainable machine learning. Among all the factors, age, BMI, and menstrual history consistently stood out as the most influential, backed by both statistical analysis and model interpretation. Despite the challenge of working with an imbalanced dataset, where most cases were already diagnosed with osteoporosis, the model maintained high levels of precision and recall, demonstrating its reliability.

What sets our approach apart is the integration of SHAP analysis, which allows users and healthcare providers to see exactly how individual health factors influence risk predictions. For example, having a history of fractures or being postmenopausal significantly increases the predicted risk, findings that closely mirror clinical understanding.

Overall, this tool supports early intervention and prevention by offering a non-invasive, cost-effective, and easy-to-use platform for osteoporosis risk screening. It’s a step forward in making personalized, proactive healthcare more accessible.

## 5. Conclusion and Future Work

This study demonstrates the potential of AI-powered models in enhancing early osteoporosis detection. OsteoInsight combines robust prediction, explainability, and ease-of-use, making it a valuable tool for both patients and practitioners.

Our findings highlight the power of interpretable machine learning in assessing osteoporosis risk with clinical nuance and actionable detail. Among the 21 input features analyzed, Menstrual History, Age, and BMI consistently emerged as the most influential predictors across both Random Forest and SHAP importance analyses. SHAP-level interpretations further revealed that specific conditions such as being postmenopausal, having a history of fractures, or reporting low estrogen signs like hot flashes or vaginal dryness, significantly increase osteoporosis risk. Conversely, factors like regular menstruation, high dairy consumption, physical activity, and calcium/vitamin D supplementation show a clear protective effect. These insights affirm established clinical knowledge while also offering granular, personalized risk explanations. Our web-based tool, built on real patient data, serves as a non-invasive, scalable solution to support early screening. For future work, we aim to expand the dataset across diverse populations, integrate BMD (Bone Mineral Density) scans for direct validation, and refine the interface for broader public and clinical use. Future work will explore additional algorithms, feature engineering (e.g., genomics or wearable data), and integration with electronic health record (EHR) systems. A longitudinal study may further validate real-world clinical outcomes.

## Data Availability

No datasets were generated or analyzed during the current study. Data sharing is not applicable to this article.

## Data and Code Availability

The source code, implementation files, and related materials for this study are publicly available on GitHub at: https://github.com/VahidMonfared/AI-Powered-Osteoporosis-Detection. The web-based research demonstration is available at: https://osteo-insight-hub.lovable.app/.

